# Joint CBC-ICT Interpretation for the pre-surgical screening of COVID 19 asymptomatic cases: A cross-sectional study

**DOI:** 10.1101/2020.07.16.20138354

**Authors:** Tanzeel Imran, Humera Altaf Naz, Hamza Khan, Ali Haider Bangash, Laraib Bakhtiar Khan

## Abstract

**Background:** On 26^th^, February 2020, first cases of COVID 19 were confirmed in Pakistan. Since then, surgeries were halted in a bid to prevent transmission. However, since such a long halt is infeasible, a general protocol of screening the carriers, especially asymptomatic carries, is a dire need of time. The objective of our study is to propose an economically feasible protocol of COVID 19 screening. Simple but effective screening strategies can help to restore the workings of hospital surgical departments.

**Methods:** We analyzed the clinical data of patients turning up for elective surgeries at the Rawal General Hospital (RGH), Islamabad from the 24^th^ of March to the 15^th^ of May, 2020. Asymptomatic patients with negative COVID 19 contact and travel histories were screened with COVID 19 Immunochromatography (ICT) IgM / IgG Ab Test. Complete blood count (CBC) was done and interpreted in conjunction with the ICT results.

**Results:** 39 patients with a mean age of 49 years were studied. The result of ICT for COVID-19 was positive in 9 cases (23%). The entire positive ICT patients population expressed significantly lower lymphocyte count (p<0.01); 8 patients had high monocyte count (p<0.05) whereas only 4 patients had a combined high neutrophil and monocyte count (P<0.05). All of these four patients with high neutrophil count were females. The combined interpretation of CBC and ICT IgM / IgG Ab Test had a high accuracy in diagnosing asymptomatic COVID-19 carriers that were later confirmed by real-time reverse transcriptase-polymerase chain reaction (rRT-PCR).

**Conclusion:** We propose that joint CBC-ICT interpretation should be adopted on a large scale to help in the diagnoses of asymptomatic carriers as both tests are simple and inexpensive and thus suit the developing countries’ limited health budget. Future research projects should be adopted in order to assess the accuracy of the proposed protocol on a large scale.

## 1 Introduction

COVID 19 pandemic, starting from a wet market in Wuhan ended up spreading globally^1^. Characteristically, patients present with fever, body aches, diarrhoea and shortness of breath that can lead, in severe cases, to pneumonia, shock and organ failure. However, a large number of patients are asymptomatic and never come under suspicion for diagnosis^2^. Complete blood count (CBC) is the most readily available and cost-effective laboratory test. If done & interpreted in conjunction with ICT antibody screening, it can help in the diagnosis of COVID 19. This being indicated, the accuracy of many antibody screening tests is being questioned by WHO^3,4^.

So far, there is no effective treatment protocol and vaccination for COVID 19 pandemic. Comprehensive support and prevention is the only option, especially for developing world owing to their limited resources. Mortality figures, as reported across the globe, are about 2%^3,5^. Elderly patients are more susceptible to complications secondary to concurrent co-morbidities. In contrast to the initial figures, significant morbidity and mortality have been reported in young adults^6^.

Routine CBC combined with the ICT IgM / IgG COVID Ab test can serve as a first-line screening test to identify asymptomatic carriers^6,7,8^. The provision of health services to non-COVID 19 patients can thus be restored^9-10^. Since the efficacy & accuracy of CBC combined with ICT IgM / IgG Ab Test interpretation to screen out COVID-19 carriers has not yet been definitely established ^11-14^, we, at RGH, aimed to evaluate the accuracy of these 2 tests in the diagnosis of asymptomatic COVID 19 cases, especially in the hospital setting before elective surgeries in order to protect paramedical staff, community and non-COVID 19 patients^15^.

## 2 Materials and methods

All of the patients planned for elective surgeries were included and their pre-surgical blood samples, as a protocol for the COVID 19 diagnosis, were collected at the RGH Pathology department, Islamabad, Pakistan. All of the patients signed a consent form approved by the RGH Ethics committee for COVID 19 ICT IgM and IgG Ab Test in conjunction with CBC test. The CBC and serology testing were done with whole blood on 39 patients. All of the patients, at the time of being received at the Pathology desk, were asymptomatic; Only 2 patients experienced mild cough. The CBC and ICT IgM / IgG serology were done within 1 hour of the collection of blood. Anti-human IgG / IgM monoclonal antibodies along with anti-goat IgG polyclonal antibodies were used in the ICT kit manufactured by Core Technology CO.LTD, China. Statistical analysis was done by SPSS version 22. Frequencies were calculated for the demographics of the study population. Chi-square test was used to determine the statistical significance of CBC and ICT IgM/IgG Ab test.

## 3 Results

In our study, we tested 39 surgical OPD patients coming in for elective surgeries. Whole blood was collected for CBC and ICT IgM / IgG Ab testing, apart from the respective routine pre-surgical investigations. A total of 9 patients were reactive on the ICT IgM / IgG Ab Test out of which only 3 patients were double-positive for IgG and IgM. 4 patients had only-IgG reactivity. Moreover, 2 patients had only-IgM reactivity. The CBC indicated mild neutrophilia (ANC) in all of the 4 female patients with count >7.99×10^3^/ul (8-19 X 10^9^ /L) whereas low normal neutrophilic count was indicated in males at 1.9 (1.9 −3.5 X 10^9^/L). Lymphopenia was featured in 100 % of the sample with absolute lymphocyte count 0.7-x10^9^/L (0.7 −0.95 X 10 ^9^/L) whereas no patient had lymphocyte account less than 0.5 X10^9^/L. While 8 patients had absolute monocyte count greater than 0.9 (1.2 −1.8 10^9^/L), the eosinophilic count was found to be within the normal range in our study population. The CBC of only 1 female patient exhibited anaemia that was attributed to menorrhagia that was confirmed by the allied test reflecting low ferritin. Thrombocytopenia was not noted in any of the COVID 19 positive asymptomatic carriers.

All of the 9 COVID 19 positive patients’ surgeries were postponed and they were quarantined at home after taking samples for the COVID 19 reverse transcription-polymerase chain reaction (RT-PCR) test. The COVID 19 RT PCR was positive in only-IgM reactive patients and combined IgG & IgM reactive patients but was negative in 1 patients (11%) out of 4 only-IgG reactive patients. The significance (p-value) for the ICT IgM/ IgG test was <0.01 whereas that of the combined interpretation of CBC and ICT IgM / IgG Ab Test parameters was <0.05. No patient was admitted and no mortality noted in the RGH study group.

## 4 Discussion

The risk associated with COVID 19 pandemic for health care workers in hospital settings can not be overstated. With the high income, developed countries offering sequencing studies, RT-PCR and Enzyme-linked immunosorbent assay (ELISA) for the community screening and diagnosis, middle & low income, developing countries, such as Pakistan is testing only symptomatic patients with RT-PCR secondary to limited resources at hand. Our findings from the RGH study group highlighted the high accuracy of collective CBC and ICT IgM / IgG Ab test interpretation in the diagnosis of asymptomatic COVID 19 cases^6,7^.Variations were detected in the CBC parameters among both; ICT-reactive & ICT non-reactive asymptomatic patients. Contrary to Mardani et al.^6^, WBC and ANC were found to be raised in ICT-reactive patients but the lymphocytic count was low. Chen et al^4^ reported similar findings in their study where they used RT-PCR for the COVID 19 diagnosis. After noting that the viral disease severity is gauged in terms of lymphocyte count, we did not find any patient with ALC <0.5 X 109/L as all were asymptomatic. Similar findings of monocytosis and lymphopenia were also reported for MERS-CoV patients in 2016.

CBC testing showed no leucopenia with normal neutrophilic count; neutrophilia was observed in 4 patients only^5,15^. Consequently, it is suggested that the neutrophils are not affected in COVID 19’s mild disease^3^ & also that SARS-CoV-2 primarily act on lymphocytes ^6-7^. ICT Antibody tests studies are reported scarcely but promising if done in conjunction with contact history, history of the symptoms of flu and diarrhoea as well as CBC^7-10^. With this indicated, it is worthwhile to indicate that in the case of a SARS-COV-2 mutation, current antibody tests detection is rendered questionable. In order to gauge the true prevalence of infections, large testing projects are required to predict true incidence. ICT is the only best solution in under-resourced, middle & low-income countries^8^.

In the present study, simple laboratory data such as CBC with differential count, when interpreted in conjunction with ICT, is found to be useful in the diagnosis of asymptomatic patients in under-resourced, developing countries where limited affordability bars the large-scale usage of RT PCR for COVID 19 diagnosis.

## 5 Limitations

Since the cohort’s size is small, the findings cannot be amounted to be general. With the study only providing a snapshot of the present, the proposed protocol should be validated by retrospective as well as prospective studies.

## 6 Conclusion

We propose that combined CBC-ICT interpretation should be tested on a large scale to diagnose asymptomatic carriers as both tests are simple and inexpensive, thus suit the developing countries that have limited health budget.

## Data Availability

The data referred to in the manuscript is maintained in a classified manner & can not be shared.

## 8 Contributor’s Statement

**Tanzeel Imran:** Conceptualization, Data curation, Investigation, Writing; **Humera Altaf Naz:** Methodology, Investigation; **Hamza Khan:** Reviewing; **Ali Haider Bangash:** Writing & Editing; **Laraib Bakhtiar Khan:** Methodology & Original draft preparation;

## 9 Acknowledgements

We are humbled to acknowledge Dr Javed and Dr Khaqan Waheed Khawaja for providing logistics support for the study. Dr Osman and Dr Haroon Khan supervised our work and guided us.

---

**Fig. 1.**
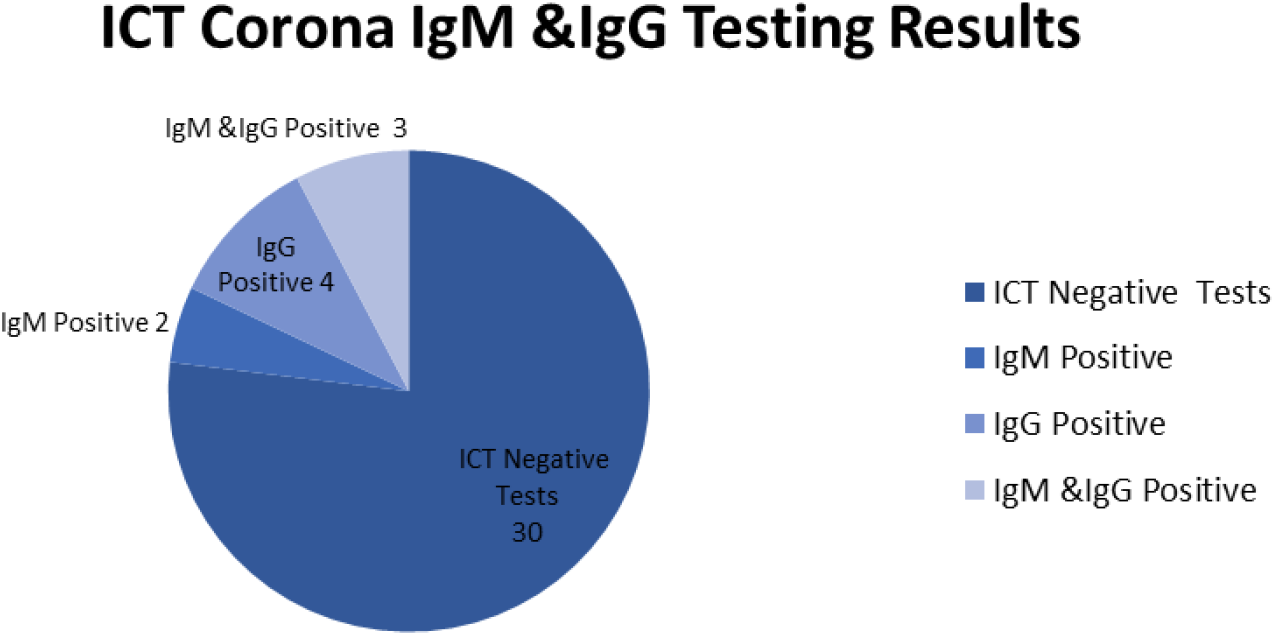
Serological detection of 2019-nCoV in asymptomatic pre-surgical screening protocol patient.

**Table 1.**
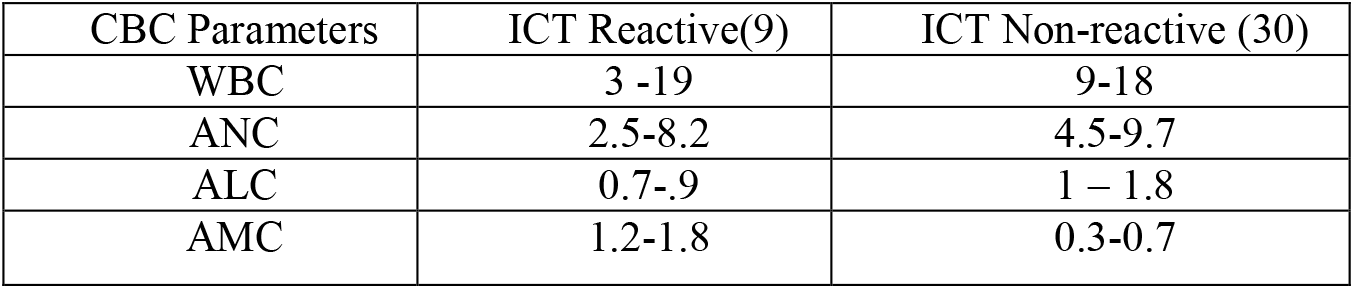
Analyzing the CBC parameters in ICT reactive COVID-19 patients.

| CBC Parameters | ICT Reactive(9) | ICT Non-reactive (30) |
| --- | --- | --- |
| WBC | 3 -19 | 9-18 |
| ANC | 2.5-8.2 | 4.5-9.7 |
| ALC | 0.7-.9 | 1 – 1.8 |
| AMC | 1.2-1.8 | 0.3-0.7 |

## Notes

### Competing Interest Statement

The authors have declared no competing interest.

### Funding Statement

The authors or their respective institutions at any time did NOT receive payment or services from a third party for any aspect of the submitted work (including but not limited to grants, data monitoring board, study design, manuscript preparation, statistical analysis, etc.).
No external funding of any kind was received.

### Author Declarations

Formal consent was obtained from the RGH Ethics committee for the COVID 19 ICT IgM / IgG Ab Test in conjunction with CBC test. Moreover, informed consent was obtained from the study participants.

### Summary of Updates

The paper has been revised only to update the list of authors.

## References

1. Li Q, Guan X, Wu P, et al. Early transmission dynamics in Wuhan, China, of novel coronavirus-infected pneumonia. N Engl J Med. 2020.

2. World Health Organization. WHO Press Statement Related to the Novel Coronavirus Situation, 2019.

3. Wang Y, Liu Y, Liu L, Wang X, Luo N, Ling L. Clinical outcome of 55 asymptomatic cases at the time of hospital admission infected with SARS-Coronavirus-2 in Shenzhen, China. The Journal of Infectious Diseases. 2020.

4. Huang C, Wang Y, Li X, et al. Clinical features of patients infected with 2019 novel coronavirus in Wuhan, China. Lancet. 2020.

5. C.L Vanessa, S Stephrene H Gek, G Kian et al. Hematologic parameters in patients with COVID-19 infection.ASH:04 March 2020

6. Mardani R, Vasmehjani A.A and Ahmad N.Laboratory Parameters in Detection of COVID-19 Patients with Positive RT-PCR; a Diagnostic Accuracy Study.. Arch Acad Emerg Med. 2020; 8(1): e43. Published online 2020 Apr 4.

7. Antibody responses to SARS-CoV-2 in patients with COVID-19. Long QX, Huang AI, et al. Nat Med. 2020 Apr 29.

8. Zhao J, Yuan Q, Wang H, et al. Antibody responses to SARS-CoV-2 in patients of novel coronavirus disease 2019. Clin Infect Dis 2020 March 28 (Epub ahead of print)

9. Arevalo-Rodriguez I, Buitrago-Garcia D, Simancas-Racines D, et al. False-negative results of initial RT-PCR assays for COVID-19: a systematic review. April 21, 2020.

10. Watson J, Whiting PF, Brush JE. Interpreting a covid-19 test result. BMJ 2020;369:m1808–m1808.

11. Wang N, Li SY, Yang XL, et al. Serological evidence of bat SARS-related coronavirus infection in humans, China. Virol Sin. 2018;33:104

12. Lei J, Li J, Li X, Qi X. CT imaging of the 2019 novel coronavirus (2019-nCoV) pneumonia. Radiology. 2020:200236.

13. Zhu N, Zhang D, Wang W, et al. A novel coronavirus from patients with pneumonia in China, 2019. N Engl J Med. 2020.

14. Zhou P, Yang XL, Wang XG, et al. Discovery of a novel coronavirus associated with the recent pneumonia outbreak in humans and its potential bat origin. BioRxiv. 2020.

15. Ding Y, He L, Zhang Q, et al. Organ distribution of severe acute respiratory syndrome (SARS) associated coronavirus (SARS-CoV) in SARS patients: implications for pathogenesis and virus transmission pathways. J Pathol. 2004;203(2):s622–630

